# Somato-cognitive action network in laryngeal and focal hand dystonia sensorimotor dysfunction

**DOI:** 10.1101/2025.02.21.25322612

**Authors:** Yuchao Wang, Baothy Huynh, Jianxun Ren, Mo Chen, Wei Zhang, Dan Hu, Shasha Li, Hesheng Liu, Teresa J. Kimberley

**Affiliations:** Center for Cognitive Neuroscience, Duke University, Durham, NC, 27708, USA; Department of Physical Therapy, MGH Institute of Health Professions, Boston, MA, 02129, USA; Department of Rehabilitation Science, MGH Institute of Health Professions, Boston, MA, 02129, USA; Changping Laboratory, Beijing, 102206, China; Neurosciences Research Program, Gillette Children’s Specialty Healthcare, St Paul, MN, 55101, USA; Non-invasive Neuromodulation Laboratory, University of Minnesota, Minneapolis, MN, 55414, USA; Academy for Advanced Interdisciplinary Studies, Peking University, Beijing, 100871, China; Department of Radiology and Biomedical Research Imaging Center, University of North Carolina at Chapel Hill, Chapel Hill, NC, 27514, USA; Athinoula A. Martinos Center for Biomedical Imaging, Department of Radiology, Massachusetts General Hospital, Boston, MA, 02129, USA; Harvard Medical School, Boston, MA, 02215, USA; Biomedical Pioneering Innovation Center, Peking University, Beijing, 100871, China

**Keywords:** neurophysiology, motor control

## Abstract

The central pathology causing idiopathic focal dystonia remains unclear, limiting effective treatment targets. The recently identified somato-cognitive action network (SCAN) with its role in coordinating physiologic processes and coarse movements has been implicated in dystonia dysfunction. SCAN is thought to interface between the phylogenetically newer primary motor regions that control fine movements and the cingulo-opercular network (CON) that putatively conveys cognitive intentions for action. We hypothesized that the effector-agnostic nature of SCAN may constitute a central pathology shared across focal dystonia subtypes affecting different body parts. Additionally, the effector-specific areas in the primary sensorimotor cortex may show distinct functional changes depending on the dystonic body region.

We collected functional MRI from patients with either of two subtypes of focal dystonia (laryngeal dystonia or LD, *N*=24, and focal hand dystonia or FHD, *N*=18) and healthy control participants (*N*=21). Regions of interest were selected based on prior work that suggested dystonia-related abnormality within the basal-ganglia-thalamo-cortical and cerebello-thalamo-cortical sensorimotor circuitries. We investigated if focal dystonia is associated with resting-state functional connectivity changes 1) between SCAN and other cortical regions (effector-specific areas and CON), 2) between cortical and non-cortical regions, or 3) between non-cortical (subcortical and cerebellar) regions. Cortical regions were individualized based on resting-state data. Separately, individualized hand and mouth/larynx regions were also generated from task-based MRI (finger-tapping and phonation, respectively) for comparison.

There was a shared interaction effect in both focal dystonia subtypes (*p*=0.048 for LD, *p*=0.017 for FHD) compared to controls, which was driven by SCAN’s higher functional connectivity to task-derived mouth/larynx region and concomitantly lower connectivity to CON. This dystonia-dependent interaction was not observed with the resting-state mouth/larynx region. No significant resting-state functional changes were observed involving subcortical and cerebellar regions when LD and FHD were modeled as independent groups. However, exploratory analysis combining LD and FHD suggested a dystonia-dependent asynchronization between SCAN and sensorimotor cerebellum (*p*=0.010) that may indicate a pathological rather than compensatory process.

For the first time, our study systematically tested circuitry-based functional connectivity changes in two focal dystonias. Our results show that SCAN is uniquely associated with dystonia dysfunction beyond the dystonic effector regions, potentially offering insights on pathophysiology and treatments.

## Introduction

A somato-cognitive action network (SCAN) involving the primary motor cortex was recently revealed through individualized functional parcellation.^1^ This network, which interdigitates between effector-specific regions of hand, foot, and mouth in the motor cortex, is hypothesized to play a key role in movement planning and coordination across sensorimotor and autonomic body control. For example, human’s evolved laryngeal control for speech may utilize SCAN to coordinate breathing. Similarly, fine hand motor control (e.g., playing a musical instrument) requires hand-eye and trunk coordination that may be mediated through SCAN. In contrast, the effector regions (hand, foot, mouth) are thought to be phylogenetically newer than SCAN, and can synapse directly with motoneurons (instead of indirectly via spinal neurons) to enable a greater range of learned and precise movements in each muscle group.^1,2^

Humans can experience discoordination and localized, abnormal muscle contractions (e.g., in their laryngeal or hand muscles) that disrupt the ability to speak, play an instrument, or write, in the absence of neurological insult or other symptoms. This disease, known as idiopathic focal dystonia, has common subtypes such as laryngeal dystonia (LD) and focal hand dystonia (FHD).^3^ Our understanding of focal dystonia has shifted from being exclusively a motor disease to a network disorder that involves sensorimotor and possibly cognitive-affective functions.^4–6^ Still, most whole-brain neuroimaging studies identify key nodes in sensorimotor pathways, specifically the basal-ganglia-thalamo-cortical and cerebello-thalamo-cortical pathways (Fig. 1A).^3,7–9^ Given that SCAN was previously not distinguished in focal dystonia, it is unclear if SCAN has differential functional connectivity with basal-ganglia and cerebellar circuitries compared to the effector-specific regions within the primary sensorimotor cortex.

**Figure 1:**
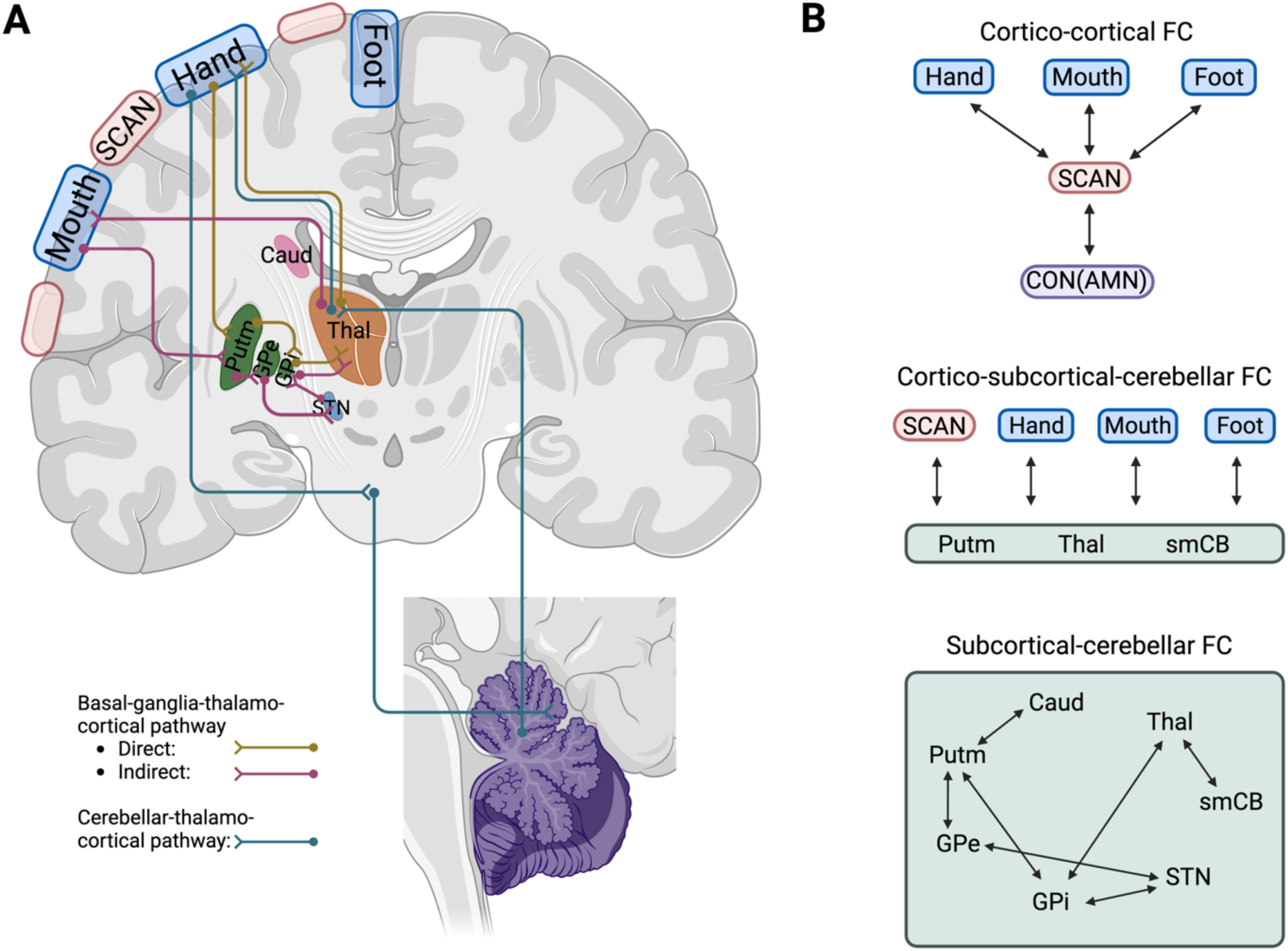
Overview of cortical-subcortical-cerebellar pathways in sensorimotor control. The recently revealed somato-cognitive action network (SCAN) may be involved in focal dystonia pathophysiology. **A)** Simplified basal-ganglia-thalamo-cortical and cerebellar-thalamo-cortical pathways are shown with the relevant subcortical structures and cerebellum. The effector-specific (hand, foot, mouth) regions are interdigitated by the SCAN in the primary motor cortex (bilaterally, with only the left hemisphere labeled). **B)** Hypothesized functional connectivity (in black arrows) that may demonstrate different SCAN vs. effector region involvement in focal dystonia affecting different body parts. We also explored connectivity within subcortical-cerebellar circuitry. FC: functional connectivity. CON: cingulo-opercular network. AMN: action-mode network. Caud: caudate. Putm: putamen. GPe: globus pallidus externus. GPi: globus pallidus internus. STN: subthalamic nucleus. Thal: thalamus. smCB: sensorimotor cerebellum. Created in BioRender.

Preliminary evidence suggests that SCAN may play an important role in focal dystonia etiology and treatment. In a pediatric population with lesion-induced dystonia, symptom-inducing lesions (in the putamen and globus pallidus) shared functional connectivity to cortical SCAN.^10^ In another study investigating where subthalamic deep brain stimulation (DBS) targets were most effective among idiopathic focal dystonia subtypes, greater symptom improvement for cervical dystonia was associated with positive functional connectivity with the cingulo-opercular network (CON),^11^ which was functionally related to SCAN. Interestingly, this SCAN-CON connectivity was not observed for appendicular (hand and foot) dystonia, whose effective DBS targets were more connected to effector-specific regions. Butenko *et al.*^11^ therefore, posited an axial-appendicular distinction for focal dystonia subtypes, where SCAN is more associated with focal dystonia affecting the mouth, neck, and trunk (axial) rather than the limbs (appendicular). This distinction, together with the involvement of SCAN however, have not been explicitly explored.

Furthermore, it remains unclear whether subtypes of focal dystonia (e.g., LD and FHD) share similar central pathologies specific to the affected body part (mouth/larynx and hand), limiting our understanding of their pathophysiology and treatment. Despite theories that the disrupted sensory feedback between the thalamus and primary somatosensory cortex is a key etiological factor,^6,12,13^ to our knowledge, no empirical work has compared the functional sensorimotor circuitry involving the effector-specific region across dystonia subtypes. Limited literature studying more than one focal dystonia subtype has used *a priori* masks of a single brain structure such as the striatum^14^ or whole-brain analysis without isolating effector-specific regions.^15–19^

With a novel dataset comprised of patients with LD or FHD and healthy controls in the current study, we tested the following hypotheses whether 1) SCAN is associated with focal dystonia pathology and 2) SCAN is more relevant for LD (axial) rather than FHD (appendicular focal dystonia).^11^ We also hypothesized that 3) focal dystonia dysfunction can be localized in sensorimotor control pathways in a dystonic-body-part-specific manner. We used functional MRI (fMRI), which has demonstrated cortico-subcortical-cerebellar functional connectivity^20,21^ following neuroanatomy.^22^ Given that LD and FHD symptoms occur only during tasks (i.e., task specificity) and that functional network boundaries may change during task,^9,23,24^ we generated individualized effector ROIs for hand and mouth (including the larynx) from both resting-state and task-based fMRI data (finger-tapping and phonation) independently for comparison. Individualized Foot ROI from resting-state data was included in the statistical model as a non-affected control. We took an exploratory approach to see whether any subcortical-cerebellar connections show dystonia-related changes (Fig. 1B).

## Materials and methods

### Participants

Sixty-three participants were enrolled in the current study, including 24 people with LD (both abductor (*N*=4) and adductor (*N*=20) type) (age=60.5±11.1, 17 females), 18 subjects with FHD (age=55.1±14.6, 6 females), and 21 healthy controls (age=53.4±12.7, 5 females). The study was approved by the University of Minnesota and Mass General Brigham Institutional Review Boards and all participants provided written informed consent according to the Declaration of Helsinki. All subjects with focal dystonia were at least 3 months away from their last botulinum toxin injection and were symptomatic during their affected tasks. Two subjects (both from the LD group) were removed from subsequent analysis due to excessive motion (mean relative motion greater than a predetermined threshold of 0.4mm during their resting-state scans). Dystonia symptom severity was measured with self-report on the Voice Handicap Index (VHI)^25^ for LD and the Arm Dystonia Disability Scale (ADDS)^26^ for FHD.

**Table 1.** Subject groups overview.

| Group | Sex | Age (years) | Symptom duration (years) | Symptom type | Resting-state scan mean relative motion (mm) | Severity |
| --- | --- | --- | --- | --- | --- | --- |
| Laryngeal dystonia ($N=24$) | Female: 17,<br>Male: 7 | $60.5\pm11.1$ | $17.2\pm10.1$ | Abductor: 4,<br>Adductor: 20 | $0.15\pm0.10$ | Voice handicap index: $61.8\pm18.1$ |
| Focal hand dystonia ($N=18$) | Female: 6,<br>Male: 12 | $55.1\pm14.6$ | $18.7\pm15.3$ | Left hand only: 6,<br>Right hand only: 9,<br>Bilateral hands: 3 | $0.14\pm0.06$ | Arm dystonia disability scale (%): $63.1\pm17.2$ |
| Control ($N=21$) | Female: 5,<br>Male: 16 | $53.4\pm12.7$ | N/A | N/A | $0.12\pm0.07$ | N/A |

### MRI acquisition

Structural and functional MRI were collected with a 3T Siemens Magnetom Prisma fit scanner with a 32-channel head coil (Siemens Healthineers, Forchheim, Germany). T1-weighted structural imaging used a 3D magnetization-prepared rapid gradient-echo imaging (MPRAGE) sequence (TR=2.5s, TI=1.0s, 0.8mm isotropic voxels, FoV=256mm, 208 sagittal slices, flip angle=8°, bandwidth=740Hz/Px). Hand and Vocal task-based fMRI were obtained with multiband gradient-echo echo-planar imaging (EPI, TR=800ms, TE=37ms, 2.0mm isotropic voxels, FoV=208mm, 72 sagittal slices, flip angle=52°, bandwidth=2290Hz/Px, echo spacing=0.58ms, 495 volumes per session). Resting-state fMRI were obtained with fast gradient-echo EPI (TR=3000ms, TE=30ms, 3.0mm isotropic voxels, FoV=216mm, 47 sagittal slices, flip angle=85°, bandwidth=2240Hz/Px, echo spacing=0.51ms, 100 volumes per session). All participants demonstrated understanding of the Hand and Vocal tasks and resting-state requirements prior to entering the MRI scanner.

For task-based scans, participants repeated the motor tasks interleaved with rest blocks in the MRI, with each block lasting 12s. Participants performed self-paced left and right index finger tapping in alternating blocks for the Hand task. For the Vocal task, participants repeated the phonation of /i:/ (“ee”) in the task block. Each task scan session lasted 396s. Participants completed one Hand task session (6.6min) and two Vocal task sessions (13.2min) to ensure a robust laryngeal activation pattern.^27^ For resting-state scans, participants were instructed to remain still with eyes closed, staying awake and relaxed without thinking of anything specific. Each resting-state scan lasted 300s and each participant completed four resting-state runs (20min total).

### MRI processing

Task-based and resting-state fMRI data were preprocessed through an identical pipeline, previously described^28,29^ and implemented in the personalized Brain Functional Sectors Cloud v1.0.7 (Neural Galaxy Inc., Beijing, China). Preprocessing included discarding the first four volumes of each session for T1 stabilization, slice timing correction via SPM12 (Wellcome Center for Human Neuroimaging, London, UK), motion correction via FSL (FMRIB, Oxford, UK), normalization of global mean signal intensity, bandpass filtering (0.01-0.08Hz), and regression of head motion and average signals from the whole brain, ventricles, and white matter. Individual cortical surfaces were registered to the *fsaverage6* template via FreeSurfer v6.0 (MGH, Boston, MA, USA). Individual subcortical and cerebellar volumes were registered to the MNI152 template (MNI152NLin6Asym) with a 2mm isotropic voxel size. A 6mm full-width-half-maximum Gaussian smoothing kernel was then applied to both surface and volumetric data to improve the signal-to-noise ratio. The residual blood-oxygen-level-dependent (BOLD) signals were used in the following analyses, performed in FreeSurfer v7.4 and custom MATLAB scripts (The MathWorks Inc., Natick, MA, USA).

### ROI definition

#### Surface task-based ROI

The motor task activation areas were used to determine the task-based ROIs (Hand and Vocal) on an individual level. First, task-related regressors (i.e., left or right finger-tapping vs. rest, or phonation vs. rest), which were convolved with a canonical hemodynamic response function, were added in the general linear model (GLM) of the subject-level task data with six parameters of motion regressor. Vertex-wise beta values associated with the motor tasks were then Z-score transformed. Task-based ROIs were defined with an arbitrary threshold at the 90^th^ percentile of Z-scores in the primary sensorimotor region (i.e., top 10% activation in the pre- and postcentral gyri and central sulcus in Destrieux *et al.*^30^), or at a Z-score > 2.33 (i.e., one-tailed vertex-wise *p*-value<0.01, uncorrected), whichever was greater. For Hand task, separate left and right finger-tapping regressors were used to define the Hand ROI on the contralateral hemisphere, while for Vocal task, a single phonation regressor determined Vocal ROI bilaterally (Fig. 2A). The resulting task-based ROIs were quality controlled based on 1) task scan mean relative motion <0.4mm (identical to the motion threshold of resting-state scan) and 2) arbitrarily >20 vertices surviving the previous Z-score thresholding in each hemisphere, to ensure sufficient data for BOLD signal averaging (Supplementary Fig. 1-2). For task ROIs that failed quality control, Hand and Mouth networks based on the individualized resting-state parcellation below were used instead to maintain power (6 subjects for Hand ROI, 7 subjects for Vocal ROI) (Supplementary Fig. 1-2).

**Figure 2:**
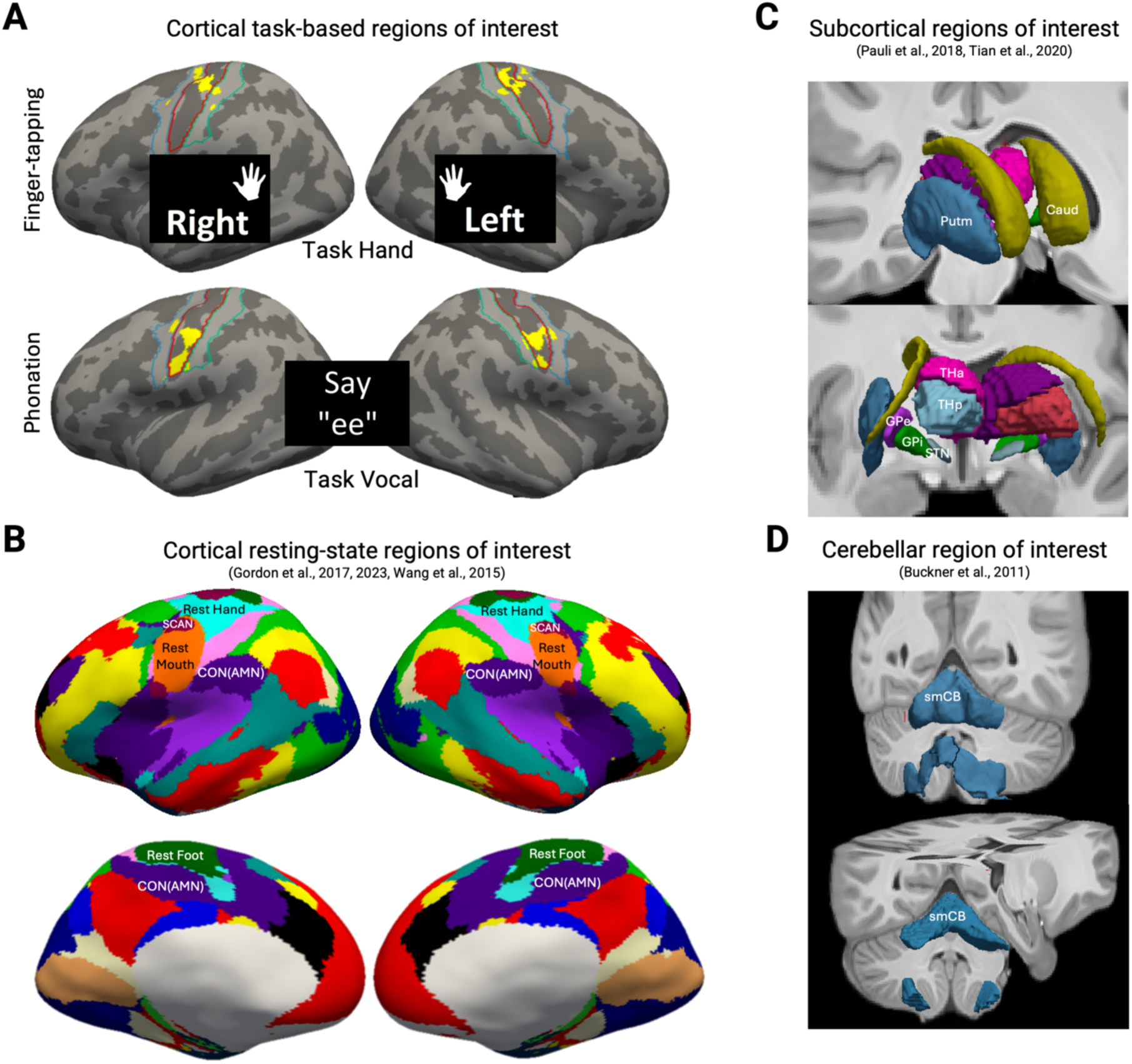
Surface and volumetric regions of interest (ROI). The affected effector regions (hand for FHD, mouth/larynx for LD) were defined from task and resting-state scans for comparison. Group atlases were used for subcortical and cerebellar volumetric ROIs. **A)** Task Hand and Task Vocal ROIs were the peak activation regions (yellow) from finger-tapping and phonation tasks, respectively, in an individualized manner (see Supplementary Fig. 1-2). Task Hand ROI on each hemisphere was separately generated from contralateral index finger tapping and combined, while bilateral Task Vocal ROI was from phonating /i:/. An *a priori* mask with pre- and postcentral gyrus (blue and tortoise outline) and central sulcus (red outline) was used to limit the task-based effector ROI to the primary sensorimotor cortex (Destrieux et al., 2010). **B)** The resting-state parcellation algorithm divided the cortex into 18 non-overlapping functional networks per hemisphere, which included Rest Hand, Rest Mouth, SCAN, CON(AMN), and (Rest) Foot. This iterative parcellation algorithm adjusted individual network boundaries based on *k*-means clustering (Supplementary Fig. 3). All surface ROIs were on the *fsaverage6* surface. **C)** Subcortical volumetric ROIs were obtained from atlases (Pauli et al., 2018, Tian et al., 2020) in MNI152 space and resampled to an isotropic voxel size of 2mm. **D)** Sensorimotor part of the cerebellum was obtained (Buckner et al., 2011) in MNI152 space and resampled to an isotropic voxel size of 2mm. SCAN: somato-cognitive action network. CON: cingulo-opercular network. AMN: action-mode network. Caud: caudate. Putm: putamen. GPe: globus pallidus externus. GPi: globus pallidus internus. STN: subthalamic nucleus. THa: anterior thalamus. THp: posterior thalamus. smCB: sensorimotor cerebellum.

#### Surface resting-state ROI

Individualized resting-state parcellations were generated using the same pipeline previously described.^29^ An 18-network group atlas was constructed by combining the 17-network parcellation from Gordon *et al.*^31^ with the SCAN reported in Gordon *et al.*^1^ (Fig. 2B). This group atlas was used to initialize an iterative *k*-means clustering algorithm that adjusted the network boundaries based on individual resting-state data, to minimize the heterogeneity within each functional network. The SCAN region was identified with an interdigitated topography along the primary motor cortex as expected. Effector regions (Rest Hand, Rest Mouth, and Foot) as well as CON, also known as the action-mode network (AMN)^32^ were also obtained (Supplementary Fig. 3).

#### Volumetric ROI

Subcortical gray matter and cerebellum ROIs (Fig. 2C, 2D) were generated based on previous atlases. Basal ganglia ROIs (caudate, putamen, globus pallidus externus (GPe), globus pallidus internus (GPi), subthalamic nucleus (STN)) were from Pauli *et al.*.^33^ Thalamus ROIs (anterior and posterior thalamus) were from Tian *et al.*.^34^ Sensorimotor network of the cerebellum was defined using the 7-network atlas from Buckner *et al.*.^20^ All volumetric atlases were conformed to the aforementioned MNI152-2mm space.

### FC definition

Given the strong functional coupling of bilateral homologs in resting-state scans, we combined all ROIs across both hemispheres. Resting-state FC was computed as the zero-lag Pearson’s correlation in the average BOLD time series between ROIs. A Fisher’s *r*-to-*z* transform was then performed for later statistical testing.

### Statistical analysis

We performed three separate linear models to investigate the interactions between dystonia group and FC between ROI pairs, accounting for covariates (i.e., age, sex, symptom duration, and mean relative motion during resting-state scan). All subsequent linear models used the formula:

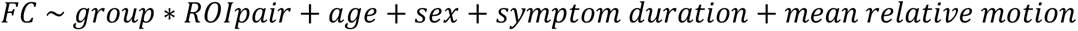

Linear model 1 tests if SCAN demonstrates dystonia-specific changes with functionally related cortical networks (effector regions and CON/AMN). Linear model 2 tests if primary sensorimotor networks (effector regions and SCAN) show dystonia-specific changes with closely synapsed subcortical and cerebellar structures (thalamus, putamen, and sensorimotor cerebellum).^1,22,35^ For linear model 1 and 2, cortical effector ROIs defined with task vs. resting-state data were included in separate models for comparison. Linear model 3 was performed to explore if any functional connection within the subcortex and cerebellum revealed dystonia-dependent changes. No multiple comparison correction was performed on the linear model estimates. Visual inspection of residuals and Breusch-Pagan tests were performed for each linear model to confirm homoscedasticity. All statistical analyses were performed in R (R Foundation for Statistical Computing, Vienna, Austria) in RStudio (Posit Software, Boston, MA, USA).

## Results

### SCAN showed dystonia-dependent interaction involving CON and task-based, but not resting-state, mouth/larynx region in both LD and FHD

There was a significant interaction effect in both LD and FHD (*p*=0.048, *p*=0.017, respectively) compared to controls, which was driven by SCAN’s higher connectivity with Task Vocal and simultaneously lower connectivity with CON. There were also main effects of CON-SCAN FC being significantly lower than those of Task Vocal-SCAN and Foot-SCAN across all groups (*p*<0.001, *p*=0.038, respectively) in the model with task-based ROI (Fig. 3B left, *F*(15,228)=7.69, *p*<0.001, *R^2^_adj_*=0.29, *p_Breusch-Pagan_*=0.083) (Supplementary Table 1). For the model with resting-state effector ROI (Fig. 3B right, *F*(15,228)=2.83, *p*<0.001, *R^2^_adj_*=0.10, *p_Breusch-Pagan_*=0.51), no dystonia-dependent interaction effect was found. There was a main effect of Foot-SCAN FC higher than CON-SCAN across all groups (*p*=0.016) (Supplementary Table 2).

**Figure 3:**
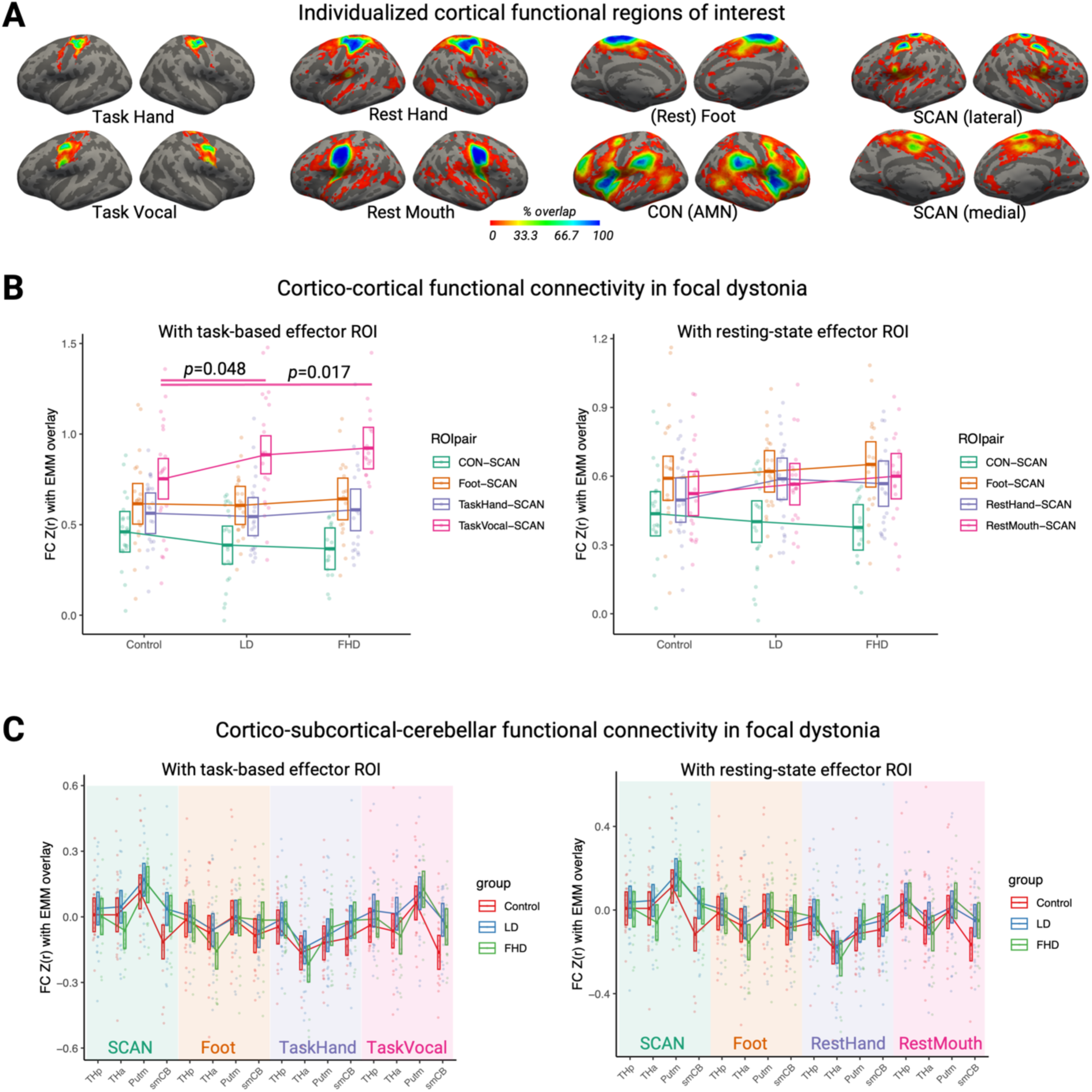
Individualized cortical ROIs and linear model results using task-based vs. resting-state effector ROIs. **A)** Individualized cortical ROIs were shown on the *fsaverage6* surface with the color scale indicating percentage overlap for all subjects in our study after quality control (*N*=57 for Task Hand, *N*=56 for Task Vocal, *N*=61 for resting-state parcellation). Foot ROI was included as an active control region. **B)** Cortical FC changes involving SCAN were tested with both task and resting-state ROIs. With task-based effector ROI (left), SCAN demonstrated a significant interaction effect for both LD and FHD groups compared to control (*p*=0.048, *p*=0.017, respectively). This interaction was driven by SCAN’s higher connectivity with Task Vocal ROI and lower connectivity with CON (i.e., AMN) in focal dystonia groups. No significant dystonia-dependent effects were found with resting-state effector ROIs (right). **C)** We tested if cortical effector regions and/or SCAN showed FC changes with closely synapsed basal-ganglia and cerebellar circuitry. Task-based and resting-state cortical ROIs were included in separate linear models. Thalamus, putamen, and sensorimotor cerebellum were included as volumetric ROIs. We did not find significant cortico-subcortical or cortical-cerebellar connections related to LD or FHD. For b-c), raw FC values after *r*-to-*z* transform were plotted with an overlay of estimated marginal mean (EMM), which accounts for covariates in the linear model (age, sex, symptom duration, and mean relative motion during resting-state scan). All error bars indicate 95% confidence interval of the EMM. SCAN: somato-cognitive action network. CON: cingulo-opercular network. AMN: action-mode network. THa: anterior thalamus. THp: posterior thalamus. Putm: putamen. smCB: sensorimotor cerebellum.

### No dystonic-body-part-specific changes were observed in basal-ganglia and cerebellar sensorimotor pathways

No significant interaction effects were found between group and ROI pairs in cortico-subcortical-cerebellar connections (Fig. 3C) or within the non-cortical regions (Fig. 4). All overall linear model fits were significant, with the greatest model explanatory power for non-cortical connectivity (Fig. 4, *R^2^_adj_*=0.72). The model details were as follows: with task-based ROI (Fig. 3C left) *F*(51,924)=4.38, *p*<0.001, *R^2^_adj_*=0.15, *p_Breusch-Pagan_*=0.098 (Supplementary Table 3); with resting-state ROI (Fig. 3C right) *F*(51,924)=4.21, *p*<0.001, *R^2^_adj_*=0.14, *p_Breusch-Pagan_*=0.37 (Supplementary Table 4); with non-cortical ROI (Fig. 4) *F*(30,518)=47.38, *p*<0.001, *R^2^_adj_*=0.72, *p_Breusch-Pagan_*=0.065 (Supplementary Table 5).

**Figure 4:**
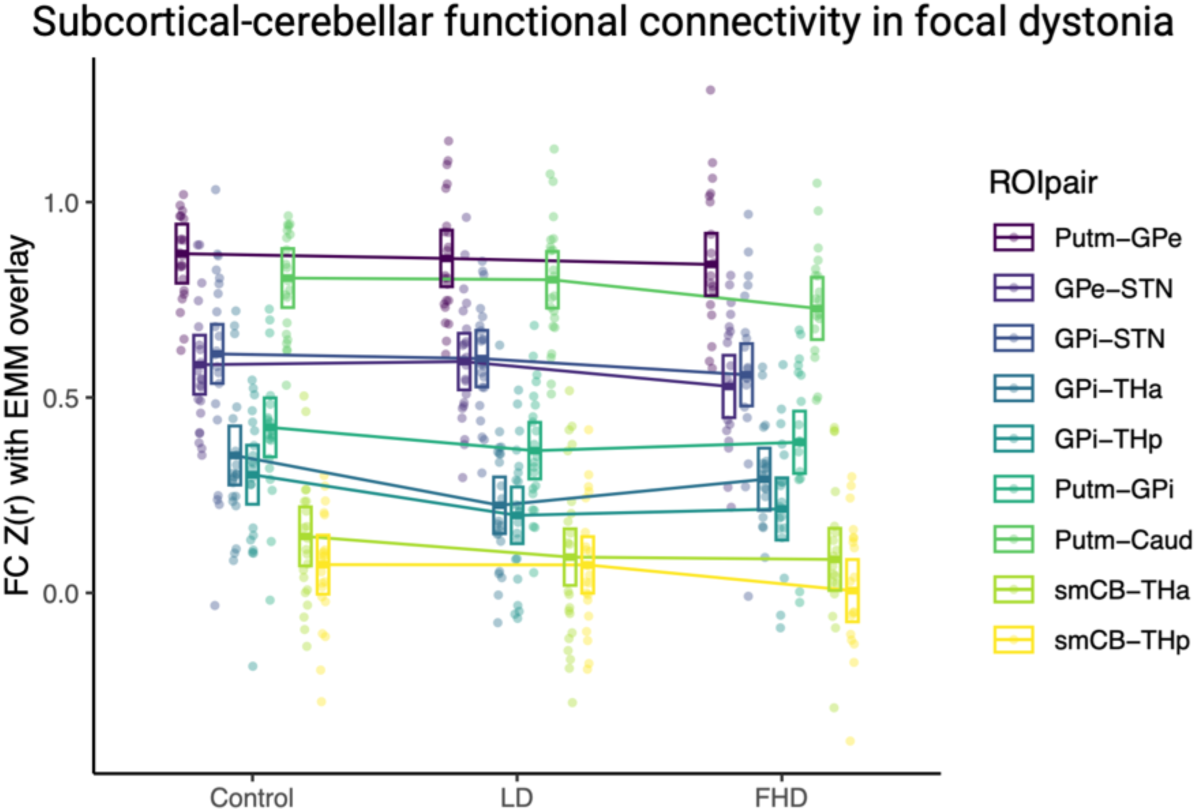
Linear model results within subcortical and cerebellar connections. No significant dystonia-dependent effects were found between *a priori* non-cortical ROIs within basal ganglia and cerebellar sensorimotor control circuitry. Raw FC values after *r*-to-*z* transform were shown with an overlay of estimated marginal mean (EMM). All error bars indicate 95% confidence interval of the EMM, which accounts for covariates in the linear model. Caud: caudate. Putm: putamen. GPe: globus pallidus externus. GPi: globus pallidus internus. STN: subthalamic nucleus. THa: anterior thalamus. THp: posterior thalamus. smCB: sensorimotor cerebellum.

### Combining LD and FHD groups suggests shared asynchronization between SCAN and sensorimotor cerebellum in focal dystonia

Given the similarity in FC distribution between LD and FHD (Fig. 3C) and the marginal interaction terms involving SCAN and sensorimotor cerebellum (*p*=0.088 for FHD, *p*=0.11 for LD, Supplementary Table 3), we combined both subtypes for increased statistical power and to explore potentially shared functional changes in focal dystonia. A *post-hoc* linear model was performed with the following formula (Fig. 5A, *F*(5,55)=2.82, *p*=0.0244, *R^2^_adj_*=0.13, *p_Breusch-Pagan_*=0.75) (Supplementary Table 6).

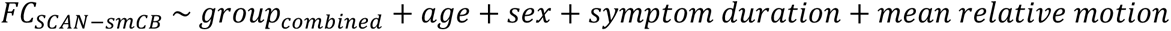

**Figure 5:**
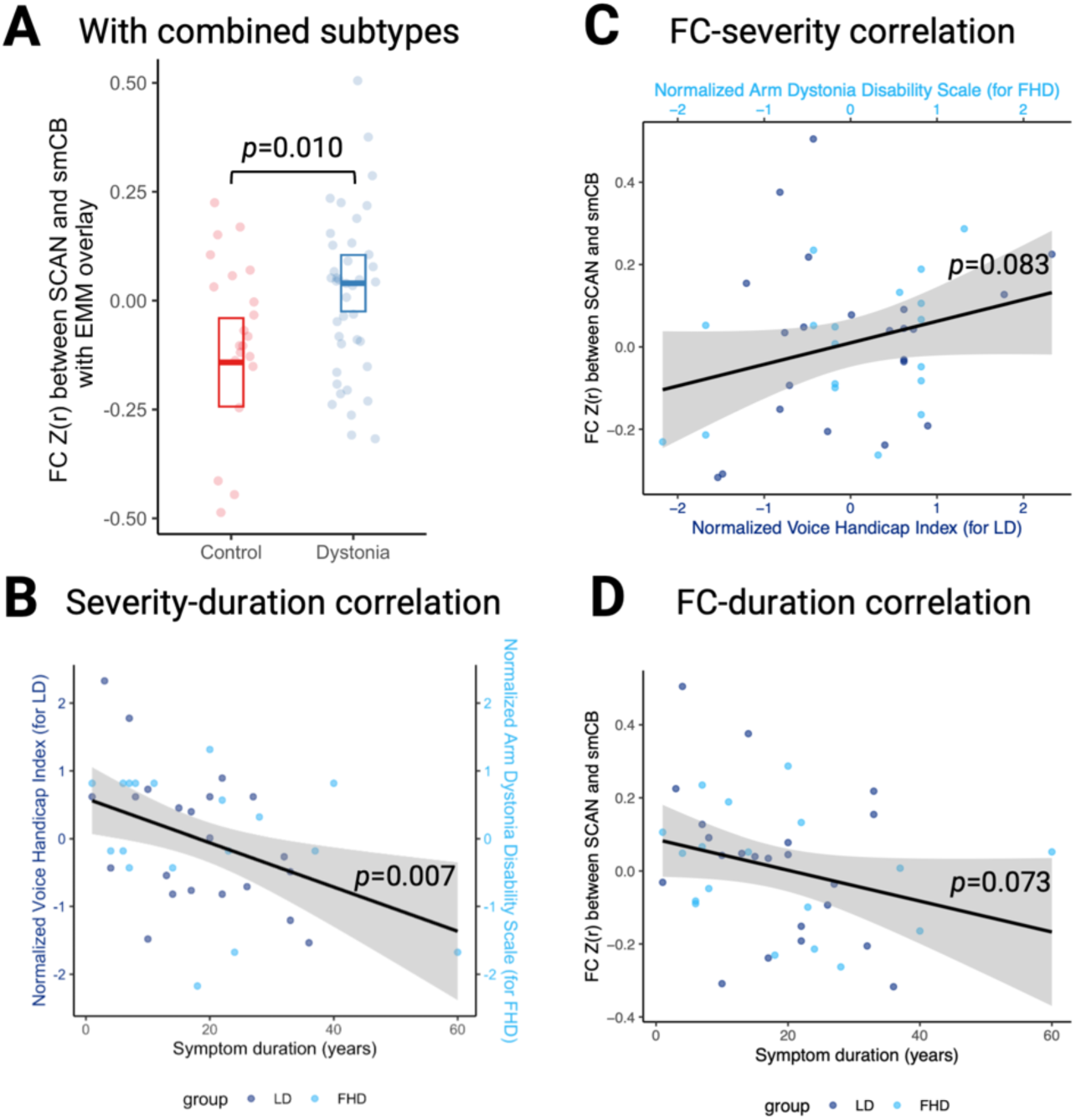
Exploratory investigation of the functional connectivity between SCAN and sensorimotor cerebellum in focal dystonia. **A)** The dystonia group combining LD and FHD showed significantly different FC (*p*=0.010) compared to controls. Specifically, there was a weak negative FC between SCAN and sensorimotor cerebellum in control subjects, which diminished to around zero in the combined dystonia group. This may suggest an asynchronization of the cortico-cerebellar sensorimotor connection with a loss of tonic inhibition at rest in focal dystonia. **B)** Dystonia severity was negatively correlated with symptom duration (*p*=0.007). Severity measures of dystonia (VHI and ADDS) were normalized (mean-centered and divided by its standard deviation) to allow being pooled for correlation tests. Higher normalized severity measure indicated greater symptoms. **C)** There was a trend towards positive correlation of SCAN-cerebellum FC against severity (*p*=0.083) and **D)** a trend towards negative correlation against symptom duration (*p*=0.073). FC: functional connectivity. EMM: estimated marginal mean. SCAN: somato-cognitive action network. smCB: sensorimotor cerebellum.

There was a significant group difference (*p*=0.010) between the combined dystonia group and controls. To further understand if this SCAN-cerebellum FC was linked to dystonia pathophysiology, we tested its correlation with symptom duration and severity in the combined dystonia group. Severity measures of dystonia (VHI and ADDS) were normalized (mean-centered and divided by its standard deviation) to allow being pooled for correlation tests. Higher normalized severity measure indicated greater symptoms. Dystonia severity was negatively correlated with symptom duration (*p*=0.007, Fig. 5B). There was a trend towards positive correlation of SCAN-cerebellum FC against severity (Fig. 5C, *p*=0.083) and a trend towards negative correlation against symptom duration (Fig. 5D, *p*=0.073).

## Discussion

For the first time, our study systematically tested if SCAN vs. dystonic effector regions demonstrate abnormal functional connectivity in cortical, basal-ganglia, and cerebellar pathways in two types of focal dystonia. Our results suggest that 1) SCAN is involved in focal dystonia pathology regardless of the affected body part (hand or larynx) and 2) dystonia-related SCAN changes may result from higher connectivity with task-based effector regions with concomitant lower connectivity with CON, and diminished negative connectivity (asynchronization) with the sensorimotor cerebellum at rest. This is consistent with our Hypothesis 1 (SCAN is associated with focal dystonia pathology) but contrary to our Hypothesis 2 (SCAN is more relevant in the axial LD rather than the appendicular FHD). There is partial support for Hypothesis 3 (focal dystonia dysfunction can be localized in sensorimotor control pathways in a dystonic-body-part-specific manner) given the group difference involving SCAN and sensorimotor cerebellum, but the functional changes were not specific to the dystonic body part.

### Individualized cortical ROIs

Accurate estimation of individual brain network variations has been a challenge especially in task-specific focal dystonia, where prior research identified topographical changes in effector networks during task^9,23,24^ perhaps due to sensorimotor reorganization.^36^ In this study, we applied a task-based ROI definition and resting-state parcellation on an individual level to compute functional connectivity in focal dystonia. The significant Task Vocal, but not Rest Mouth, dystonia-related changes may suggest that the individual variability in motor tasks is important for focal dystonia pathology and cannot be fully captured in resting-state scans. Alternatively, the laryngeal motor network (see discussion below) derived from our phonation task may be too fine-grained for accurate estimation from only resting-state data. Our results thus demonstrate the feasibility and potentially also the necessity of using individual task ROIs for focal dystonia research.

To the point above, we observed greater individual variability (i.e., less percentage overlap) in the task-derived ROIs for hand and mouth/larynx, despite similar network location on the group level compared to resting-state ROIs (Fig. 3A). Notably, our phonation task generated a ventral and dorsal pattern grossly within the Rest Mouth network, consistent with the laryngeal motor network (cf. ^37^).^1,27,38^ Our Task Vocal ROI also consisted of an area in the superior sensorimotor cortex bilaterally (left hemisphere (peak 18% overlap) at (−18.1, −24.9, 61.7) in RAS coordinates on the *fsaverage6* surface, same below; right hemisphere (peak 29% overlap) at (19.2, −25.4, 57.5)). This region was previously thought to be the trunk or abdominal area,^27,38^ but overlapped with SCAN, which likely reflected the involvement of SCAN in controlling breathing for vocalizing.^1^ Despite the partial overlap, SCAN and Task Vocal ROI are distinct functional networks, with the Task Vocal (laryngeal) ROI being located more inferiorly and anteriorly to the SCAN in the ventral primary sensorimotor cortex (for SCAN: left hemisphere (peak 98% overlap) at (−35.0, −15.1, 42.0), right hemisphere (peak 94% overlap) at (36.7, −12.7, 40.3); for Task Vocal: left hemisphere (peak 82% overlap) at (−50.0, −9.5, 45.2), right hemisphere (peak 73% overlap) at (50.5,-5.7, 45.0)).

### Cortical functional changes

One crucial unresolved question in focal dystonia is whether the pathophysiology differs depending on the body part affected.^39^ Relatively few empirical studies have included focal dystonia affecting different effectors, mostly with network analysis and graph theory, and the results have been mixed. For example, compared to healthy controls, Battistella *et al.*^16^ found commonly gained hubs in left insula and bilateral auditory cortex in LD and FHD using resting-state functional data, whereas Hanekemp and Simonyan^19^ found lost hubs in left insula but gained hub in right superior frontal gyrus with diffusion data. Also, with resting-state fMRI in LD and FHD, Fuertinger and Simonyan^18^ found widespread lost and gained hubs across sensorimotor cortex, cuneus, parietal cortex, occipital cortex, and cerebellum while Bianchi *et al.*^15^ found increased connectivity at right inferior parietal lobule. Our novel approach probing the basal-ganglia-thalamo-cortical and cerebellar-thalamo-cortical pathways in human focal dystonia may complement graph theory and ROI-based methods (e.g., ^14,36^) to bridge with extensive circuitry-level findings from animal literature.

The cortico-cortical abnormality involving the mouth/larynx region (higher SCAN-Task Vocal connectivity with lower SCAN-CON connectivity compared to controls) shared in LD and FHD (Fig. 3B) may suggest a dystonia-related predilection rather than a symptom-causing biomarker, since FHD subjects did not experience laryngeal symptoms. Clinically, it is known that focal dystonia affecting one body part may spread to other isolated body parts,^40^ pointing to common central pathology beyond the involved muscles. Given the role of the cingulo-opercular action-mode network in goal-directed actions^32^ and SCAN in mediating cognition and body control, the decreasing FC between SCAN and CON may indicate greater difficulty forming a motor plan in a top-down manner in focal dystonia.

A potential confound to our cortico-cortical interaction effect (Fig. 3B) is that Task Vocal ROI may overlap with SCAN to different extents in dystonia and control subjects, thus inflating the functional connectivity between Task Vocal and SCAN in a dystonia-dependent manner. This is unlikely to fully account for the interaction effect, however, since the overlap was low (group mean overlap <15% of the individual SCAN area) and no significant group difference was found (Supplementary Fig. 4). Additionally, the positive findings were reinforced given that the FC changes were not observed in our non-affected control ROI (Foot).

Several reasons may explain why we did not find similar dystonia-related changes in the hand region. First, unlike laryngeal control which is bilateral through the corticobulbar tract, hand control is heavily lateralized via the corticospinal tract, and its abnormal activity may therefore be hard to detect in our BOLD time courses averaged over bilateral ROIs. Second, our phonation task may be more symptom-inducing than finger-tapping, leading to more accurate estimation of the affected functional network in LD rather than FHD. Third, this task quality difference may be compounded by greater heterogeneity in FHD phenotypes, since our subjects had different hand (left, right, bilateral) and finger/wrist involvement.

### Cerebellar functional changes

Finding key pathological functional alterations can update our disease categorization,^5^ and inform potentially unifying treatment approaches that allow for reconfiguration of the pathological brain network for symptom relief.^41–43^ The SCAN-cerebellum connection in our exploratory model combining LD and FHD groups may be such a candidate (Fig. 5A). Crucially, there was a weak negative functional connectivity between SCAN and sensorimotor cerebellum in healthy controls, which diminished to around zero (indicating asynchronization) in the combined dystonia group based on the estimated marginal mean.

Similar resting-state functional changes have been reported in cervical dystonia and blepharospasm, where Giannì *et al.* observed a “loss of anticorrelation” in the primary sensorimotor cortex with a seed at the output nucleus of the cerebellum (dentate).^44^ Our results thus expand the validity of this functional asynchronization to focal dystonia affecting other body regions and distinguish the unique role of SCAN within the primary sensorimotor cortex.

To investigate if this SCAN-cerebellum connectivity may be relevant for pathophysiology in our cohort, we reasoned that the extent of the FC changes *away* from the normal (weak negative FC) should positively correlate with symptom severity. This was weakly supported by the correlation result (Fig. 5C). Alternatively, symptom duration was negatively correlated with disease severity (Fig. 5B), possibly suggesting that over time, the brain may have undergone neuroplastic changes that ameliorated the pathology. This amelioration of dystonia symptoms was marginally associated with lowered SCAN-cerebellum FC (i.e., a return to weak negative connectivity similar to controls) (Fig. 5D). It is therefore probable that SCAN-cerebellum asynchronization reflects pathological rather than compensatory changes.

The weak negative connectivity between cerebellum and SCAN may thus be a form of healthy tonic inhibition at rest, which was disrupted with focal dystonia onset. This loss of inhibition in cerebellar-thalamo-cortical pathway was consistent with prior findings mainly in FHD.^45–47^ Our results expanded the scope to include LD and isolated the unique role of SCAN cortically. Interestingly, a recent pre-clinical study found that this cerebellar-thalamo-cortical pathway was responsible for learned, context-dependent movement initiation that may offer insight to task specificity in focal dystonia.^48^ Our findings may provide support for novel treatment targets such as the cerebellum, in the context of emergent clinical trials.^49^

### Limitations

There are limitations in the current study. Our limited sample size without power analysis may lead to reduced statistical power that failed to detect functional changes e.g., in basal ganglia pathways. The self-reported disease severity measures had poor sensitivity and we did not obtain dystonia-related genetic information,^3^ which could introduce heterogeneity in our groups. Given our relatively small sample size, we also did not differentiate between different phenotypes within LD (adductor vs. abductor) and FHD (handedness, finger/wrist involvement). In combining bilateral ROIs, we omitted laterality which may be relevant even for LD.^50^ Our results on SCAN-cerebellum connectivity across LD and FHD remain correlational and need to be reconciled with more causal findings from DBS sweet spot connectivity mapping.^11^ Finally, our volumetric ROIs may be too coarse. For example, Gordon *et al.*^1^ found the centromedian nucleus (CM) of the thalamus to be functionally connected to SCAN. Due to CM’s small size, the effect would likely have been washed out by the anterior/posterior thalamic ROI segmentation.

Focal dystonia is a debilitating disease with unclear pathophysiology and limited treatment. The role of SCAN in coordinating between cognitive intention and motor execution may be particularly relevant for task-specific focal dystonia. Our results show that SCAN is uniquely associated with dystonia dysfunction beyond the dystonic effector regions, potentially offering new insights on pathophysiology and treatments.

## Data availability

Data and code used for statistical analysis and visualization are available at https://osf.io/nmdka/.

## Acknowledgements

We would like to thank Mark Hallett, Andreas Horn, and Kristina Simonyan for helpful discussions on earlier results. We thank all participants for their time and contribution to this study.

## References

1. Gordon EM, Chauvin RJ, Van AN, et al. A somato-cognitive action network alternates with effector regions in motor cortex. Nature. 2023;617(7960):351–359. doi:10.1038/s41586-023-05964-2

2. Rathelot JA, Strick PL. Subdivisions of primary motor cortex based on cortico-motoneuronal cells. Proc Natl Acad Sci U S A. 2009;106(3):918–923. doi:10.1073/pnas.0808362106

3. Balint B, Mencacci NE, Valente EM, et al. Dystonia. Nat Rev Dis Primer. 2018;4(1):25. doi:10.1038/s41572-018-0023-6

4. Rothwell JC, Obeso JA, Day BL, Marsden CD. Pathophysiology of dystonias. Adv Neurol. 1983;39:851–863.

5. Simonyan K, Barkmeier-Kraemer J, Blitzer A, et al. Laryngeal Dystonia: Multidisciplinary Update on Terminology, Pathophysiology, and Research Priorities. Neurology. 2021;96(21):989–1001. doi:10.1212/WNL.0000000000011922

6. Prudente CN, Hess EJ, Jinnah HA. Dystonia as a network disorder: What is the role of the cerebellum? Neuroscience. 2014;260:23–35. doi:10.1016/j.neuroscience.2013.11.062

7. Zoons E, Booij J, Nederveen AJ, Dijk JM, Tijssen MAJ. Structural, functional and molecular imaging of the brain in primary focal dystonia—A review. NeuroImage. 2011;56(3):1011–1020. doi:10.1016/j.neuroimage.2011.02.045

8. Kshatriya N, Battistella G, Simonyan K. Structural and functional brain alterations in laryngeal dystonia: A coordinate-based activation likelihood estimation meta-analysis. Hum Brain Mapp. 2024;45(14):e70000. doi:10.1002/hbm.70000

9. Gallea C, Horovitz SG, ’Ali Najee-Ullah M, Hallett M. Impairment of a parieto-premotor network specialized for handwriting in writer’s cramp. Hum Brain Mapp. 2016;37(12):4363–4375. doi:10.1002/hbm.23315

10. Gelineau-Morel R, Dlamini N, Bruss J, et al. Network localization of pediatric lesion-induced dystonia. Published online April 8, 2024. doi:10.1101/2024.04.06.24305421

11. Butenko K, Neudorfer C, Dembek TA, et al. Engaging dystonia networks with subthalamic stimulation. Proc Natl Acad Sci U S A. 2025;122(2):e2417617122. doi:10.1073/pnas.2417617122

12. Perruchoud D, Murray MM, Lefebvre J, Ionta S. Focal dystonia and the Sensory-Motor Integrative Loop for Enacting (SMILE). Front Hum Neurosci. 2014;8. doi:10.3389/fnhum.2014.00458

13. Conte A, Defazio G, Hallett M, Fabbrini G, Berardelli A. The role of sensory information in the pathophysiology of focal dystonias. Nat Rev Neurol. 2019;15(4):224–233. doi:10.1038/s41582-019-0137-9

14. Simonyan K, Cho H, Hamzehei Sichani A, Rubien-Thomas E, Hallett M. The direct basal ganglia pathway is hyperfunctional in focal dystonia. Brain. 2017;140(12):3179–3190. doi:10.1093/brain/awx263

15. Bianchi S, Fuertinger S, Huddleston H, Frucht SJ, Simonyan K. Functional and structural neural bases of task specificity in isolated focal dystonia. Mov Disord. 2019;34(4):555–563. doi:10.1002/mds.27649

16. Battistella G, Termsarasab P, Ramdhani RA, Fuertinger S, Simonyan K. Isolated Focal Dystonia as a Disorder of Large-Scale Functional Networks. Cereb Cortex. Published online December 17, 2015:bhv313. doi:10.1093/cercor/bhv313

17. Berman BD, Honce JM, Shelton E, Sillau SH, Nagae LM. Isolated focal dystonia phenotypes are associated with distinct patterns of altered microstructure. NeuroImage Clin. 2018;19:805–812. doi:10.1016/j.nicl.2018.06.004

18. Fuertinger S, Simonyan K. Task-specificity in focal dystonia is shaped by aberrant diversity of a functional network kernel. Mov Disord. 2018;33(12):1918–1927. doi:10.1002/mds.97

19. Hanekamp S, Simonyan K. The large-scale structural connectome of task-specific focal dystonia. Hum Brain Mapp. 2020;41(12):3253–3265. doi:10.1002/hbm.25012

20. Buckner RL, Krienen FM, Castellanos A, Diaz JC, Yeo BTT. The organization of the human cerebellum estimated by intrinsic functional connectivity. J Neurophysiol. 2011;106(5):2322–2345. doi:10.1152/jn.00339.2011

21. Ji JL, Spronk M, Kulkarni K, Repovš G, Anticevic A, Cole MW. Mapping the human brain’s cortical-subcortical functional network organization. NeuroImage. 2019;185:35–57. doi:10.1016/j.neuroimage.2018.10.006

22. Bostan AC, Strick PL. The basal ganglia and the cerebellum: nodes in an integrated network. Nat Rev Neurosci. 2018;19(6):338–350. doi:10.1038/s41583-018-0002-7

23. Simonyan K, Ludlow CL. Abnormal Activation of the Primary Somatosensory Cortex in Spasmodic Dysphonia: An fMRI Study. Cereb Cortex. 2010;20(11):2749–2759. doi:10.1093/cercor/bhq023

24. Byl NN, McKenzie A, Nagarajan SS. Differences in somatosensory hand organization in a healthy flutist and a flutist with focal hand dystonia. J Hand Ther. 2000;13(4):302–309. doi:10.1016/S0894-1130(00)80022-8

25. Jacobson BH, Johnson A, Grywalski C, et al. The Voice Handicap Index (VHI). Am J Speech Lang Pathol. 1997;6(3):66–70. doi:10.1044/1058-0360.0603.66

26. Fahn S. Assessment of the primary dystonias. In: Munsat TL, ed. Quantification of Neurological Deficit. 1st edition. Butterworth-Heinemann; 1989:241–270.

27. Chen M, Summers RLS, Prudente CN, et al. Transcranial magnetic stimulation and functional magnet resonance imaging evaluation of adductor spasmodic dysphonia during phonation. Brain Stimulat. 2020;13(3):908–915. doi:10.1016/j.brs.2020.03.003

28. Yeo BT, Krienen FM, Sepulcre J, et al. The organization of the human cerebral cortex estimated by intrinsic functional connectivity. J Neurophysiol. 2011;106(3):1125–1165. doi:10.1152/jn.00338.2011

29. Wang D, Buckner RL, Fox MD, et al. Parcellating Cortical Functional Networks in Individuals. Nat Neurosci. 2015;18(12):1853–1860. doi:10.1038/nn.4164

30. Destrieux C, Fischl B, Dale A, Halgren E. Automatic parcellation of human cortical gyri and sulci using standard anatomical nomenclature. NeuroImage. 2010;53(1):1–15. doi:10.1016/j.neuroimage.2010.06.010

31. Gordon EM, Laumann TO, Gilmore AW, et al. Precision Functional Mapping of Individual Human Brains. Neuron. 2017;95(4):791–807.e7. doi:10.1016/j.neuron.2017.07.011

32. Dosenbach NUF, Raichle ME, Gordon EM. The brain’s action-mode network. Nat Rev Neurosci. Published online January 2, 2025:1–11. doi:10.1038/s41583-024-00895-x

33. Pauli WM, Nili AN, Tyszka JM. A high-resolution probabilistic in vivo atlas of human subcortical brain nuclei. Sci Data. 2018;5(1):180063. doi:10.1038/sdata.2018.63

34. Tian Y, Margulies DS, Breakspear M, Zalesky A. Topographic organization of the human subcortex unveiled with functional connectivity gradients. Nat Neurosci. 2020;23(11):1421–1432. doi:10.1038/s41593-020-00711-6

35. Kaji R, Bhatia K, Graybiel AM. Pathogenesis of dystonia: is it of cerebellar or basal ganglia origin? J Neurol Neurosurg Psychiatry. 2018;89(5):488–492. doi:10.1136/jnnp-2017-316250

36. Kimberley TJ, Pickett KA. Differential activation in the primary motor cortex during individual digit movement in focal hand dystonia vs. healthy. Restor Neurol Neurosci. 2012;30(3):247–254.

37. Simonyan K, Ostuni J, Ludlow CL, Horwitz B. Functional But Not Structural Networks of the Human Laryngeal Motor Cortex Show Left Hemispheric Lateralization during Syllable But Not Breathing Production. J Neurosci. 2009;29(47):14912–14923. doi:10.1523/JNEUROSCI.4897-09.2009

38. Correia JM, Caballero-Gaudes C, Guediche S, Carreiras M. Phonatory and articulatory representations of speech production in cortical and subcortical fMRI responses. Sci Rep. 2020;10(1):4529. doi:10.1038/s41598-020-61435-y

39. Lungu C, Ozelius L, Standaert D, et al. Defining research priorities in dystonia. Neurology. 2020;94(12):526–537. doi:10.1212/WNL.0000000000009140

40. Defazio G, Berardelli A, Hallett M. Do primary adult-onset focal dystonias share aetiological factors? Brain. 2007;130(5):1183–1193. doi:10.1093/brain/awl355

41. Kimberley TJ, Borich MR, Arora S, Siebner HR. Multiple sessions of low-frequency repetitive transcranial magnetic stimulation in focal hand dystonia: clinical and physiological effects. Restor Neurol Neurosci. 2013;31(5):533–542. doi:10.3233/RNN-120259

42. Horn A, Reich MM, Ewert S, et al. Optimal deep brain stimulation sites and networks for cervical vs. generalized dystonia. Proc Natl Acad Sci. 2022;119(14):e2114985119. doi:10.1073/pnas.2114985119

43. Okromelidze L, Tsuboi T, Eisinger RS, et al. Functional and Structural Connectivity Patterns Associated with Clinical Outcomes in Deep Brain Stimulation of the Globus Pallidus Internus for Generalized Dystonia. Am J Neuroradiol. 2020;41(3):508–514. doi:10.3174/ajnr.A6429

44. Giannì C, Pasqua G, Ferrazzano G, et al. Focal Dystonia: Functional Connectivity Changes in Cerebellar-Basal Ganglia-Cortical Circuit and Preserved Global Functional Architecture. Neurology. 2022;98(14). doi:10.1212/WNL.0000000000200022

45. Shakkottai VG, Batla A, Bhatia K, et al. Current Opinions and Areas of Consensus on the Role of the Cerebellum in Dystonia. The Cerebellum. 2017;16(2):577–594. doi:10.1007/s12311-016-0825-6

46. Argyelan M, Carbon M, Niethammer M, et al. Cerebellothalamocortical Connectivity Regulates Penetrance in Dystonia. J Neurosci. 2009;29(31):9740–9747. doi:10.1523/JNEUROSCI.2300-09.2009

47. Meunier S, Russmann H, Shamim E, Lamy J, Hallett M. Plasticity of cortical inhibition in dystonia is impaired after motor learning and paired-associative stimulation. Eur J Neurosci. 2012;35(6):975–986. doi:10.1111/j.1460-9568.2012.08034.x

48. Dacre J, Colligan M, Clarke T, et al. A cerebellar-thalamocortical pathway drives behavioral context-dependent movement initiation. Neuron. 2021;109(14):2326–2338.e8. doi:10.1016/j.neuron.2021.05.016

49. Fan H, Zheng Z, Yin Z, Zhang J, Lu G. Deep Brain Stimulation Treating Dystonia: A Systematic Review of Targets, Body Distributions and Etiology Classifications. Front Hum Neurosci. 2021;15:757579. doi:10.3389/fnhum.2021.757579

50. Krüger MT, Hu A, Honey CR. Deep Brain Stimulation for Spasmodic Dysphonia: A Blinded Comparison of Unilateral and Bilateral Stimulation in Two Patients. Stereotact Funct Neurosurg. 2020;98(3):200–205. doi:10.1159/000507058

