## Supplementary Material for "Somato-cognitive action network in laryngeal and focal hand dystonia sensorimotor dysfunction"

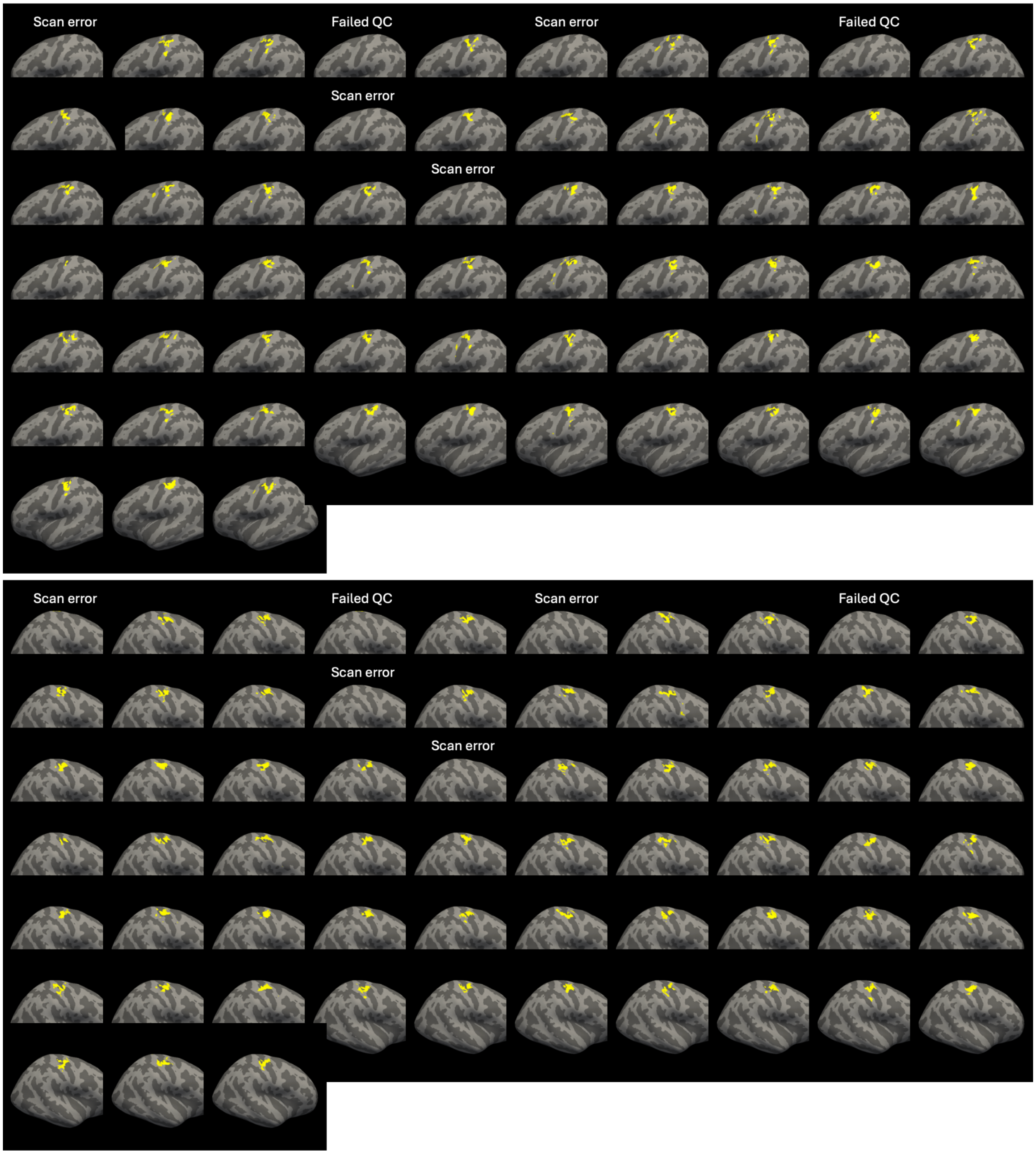


**Supplementary Figure 1: Individualized Task Hand ROI.**

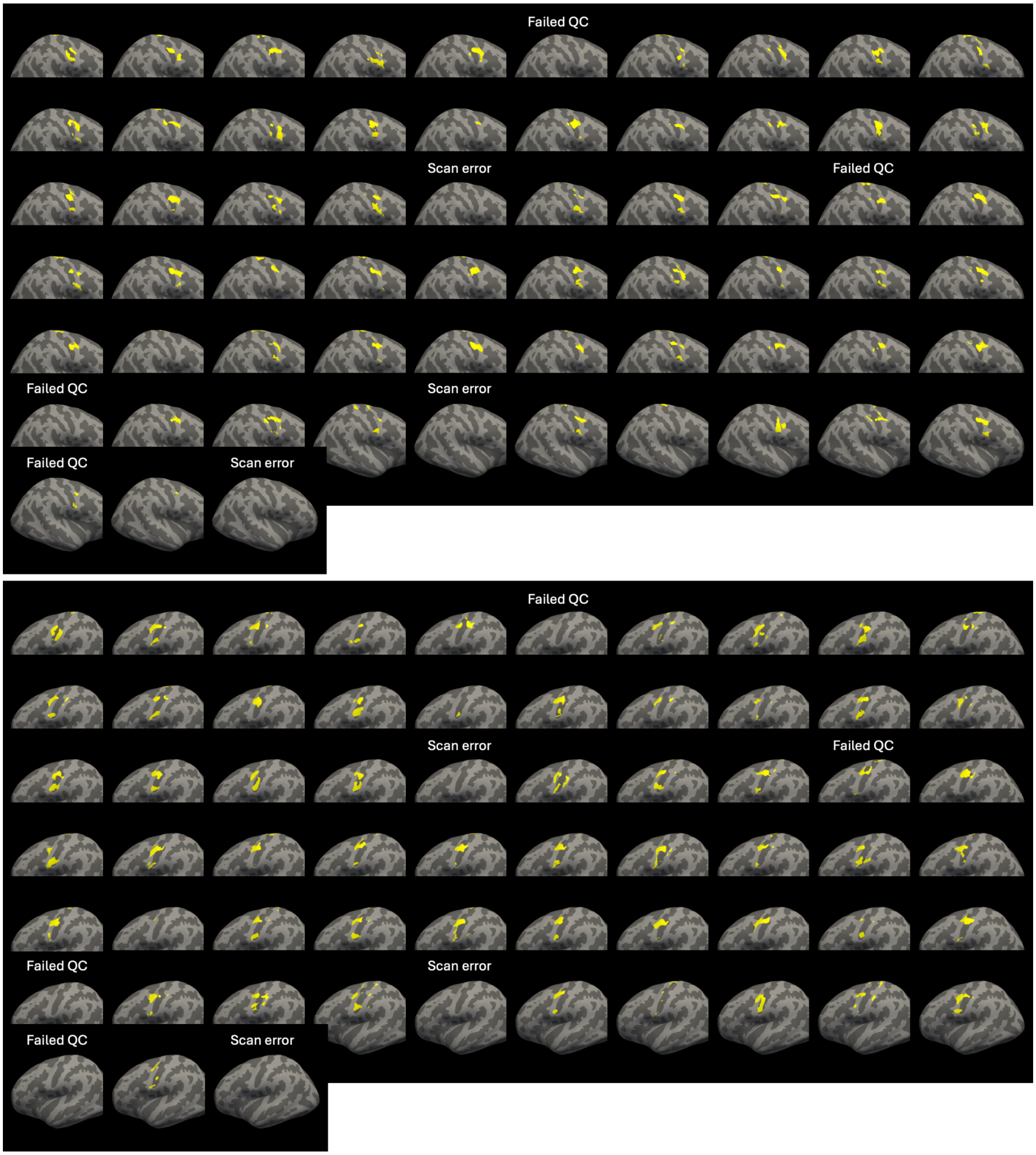


**Supplementary Figure 2: Individualized Task Vocal ROI.**

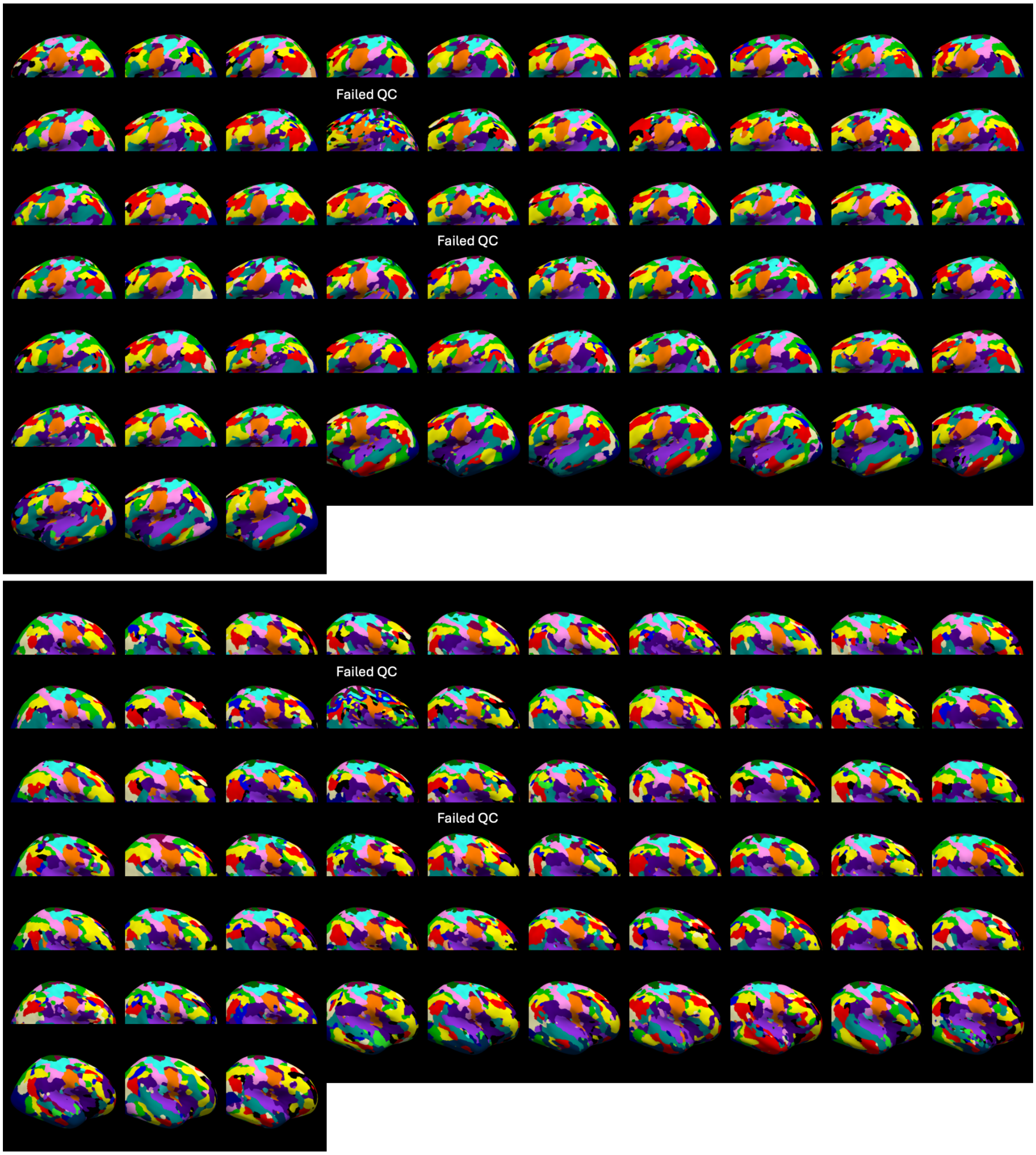


**Supplementary Figure 3: Individualized resting-state parcellation results.** Rest Hand is in cyan, Rest Mouth in orange, Foot in dark green, SCAN in marron, and CON(AMN) in dark purple. SCAN: somato-cognitive action network. CON: cingulo-opercular network. AMN: action-mode network.

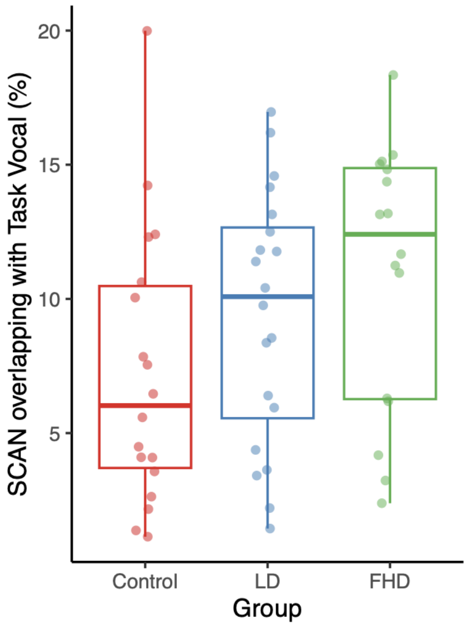


**Supplementary Figure 4: Overlap between SCAN and Task Vocal.** No significant group difference was found (one-way ANOVA, $F(2,51)=2.44$, $p=0.098$) though a trend was observed with greater overlap between SCAN and Task Vocal in focal dystonia groups.

### Supplementary Table 1: Cortico-cortical FC (linear model 1) with task-based effector ROI

| | FC | | |
|---|---|---|---|
| *Predictors* | *Estimates* | *CI* | *P* |
| (Intercept) | 0.42 | 0.23 – 0.60 | **<0.001** |
| group [LD] | -0.07 | -0.23 – 0.09 | 0.368 |
| group [FHD] | -0.09 | -0.26 – 0.07 | 0.264 |
| ROIpair [Foot-SCAN] | 0.15 | 0.01 – 0.30 | **0.038** |
| ROIpair [TaskHand-SCAN] | 0.10 | -0.04 – 0.25 | 0.170 |
| ROIpair [TaskVocal-SCAN] | 0.29 | 0.15 – 0.44 | **<0.001** |
| age | 0.00 | -0.00 – 0.00 | 0.474 |
| sex [M] | 0.01 | -0.06 – 0.08 | 0.860 |
| symptom duration | -0.00 | -0.00 – 0.00 | 0.742 |
| Mean Relative Motion | -0.07 | -0.59 – 0.45 | 0.799 |
| group [LD] × ROIpair [Foot-SCAN] | 0.06 | -0.14 – 0.27 | 0.536 |
| group [FHD] × ROIpair [Foot-SCAN] | 0.12 | -0.10 – 0.34 | 0.273 |
| group [LD] × ROIpair [TaskHand-SCAN] | 0.06 | -0.15 – 0.26 | 0.596 |
| group [FHD] × ROIpair [TaskHand-SCAN] | 0.11 | -0.10 – 0.33 | 0.305 |
| group [LD] × ROIpair [TaskVocal-SCAN] | 0.21 | 0.00 – 0.41 | **0.048** |
| group [FHD] × ROIpair [TaskVocal-SCAN] | 0.26 | 0.05 – 0.48 | **0.017** |
| Observations | 244 | | |
| $R^2$ / $R^2$ adjusted | 0.336 / 0.292 | | |

### Supplementary Table 2: Cortico-cortical FC (linear model 1) with resting-state effector ROI

| | FC | | |
|---|---|---|---|
| *Predictors* | *Estimates* | *CI* | *p* |
| (Intercept) | 0.48 | 0.32 – 0.65 | **<0.001** |
| group [LD] | -0.03 | -0.17 – 0.10 | 0.626 |
| group [FHD] | -0.06 | -0.20 – 0.08 | 0.409 |
| ROIpair [Foot-SCAN] | 0.15 | 0.03 – 0.28 | **0.016** |
| ROIpair [RestHand-SCAN] | 0.06 | -0.07 – 0.19 | 0.352 |
| ROIpair [RestMouth-SCAN] | 0.09 | -0.04 – 0.21 | 0.170 |
| age | 0.00 | -0.00 – 0.00 | 0.999 |
| sex [M] | 0.01 | -0.05 – 0.07 | 0.711 |
| symptom duration | -0.00 | -0.00 – 0.00 | 0.152 |
| Mean Relative Motion | -0.24 | -0.69 – 0.21 | 0.301 |
| group [LD] × ROIpair [Foot-SCAN] | 0.06 | -0.11 – 0.24 | 0.472 |
| group [FHD] × ROIpair [Foot-SCAN] | 0.12 | -0.07 – 0.31 | 0.204 |
| group [LD] × ROIpair [RestHand-SCAN] | 0.13 | -0.05 – 0.30 | 0.157 |
| group [FHD] × ROIpair [RestHand-SCAN] | 0.13 | -0.05 – 0.32 | 0.165 |
| group [LD] × ROIpair [RestMouth-SCAN] | 0.08 | -0.10 – 0.25 | 0.401 |
| group [FHD] × ROIpair [RestMouth-SCAN] | 0.14 | -0.05 – 0.32 | 0.152 |
| Observations | 244 | | |
| $R^2$ / $R^2$ adjusted | 0.157 / 0.102 | | |

**Supplementary Table 3: Cortico-subcortical-cerebellar FC (linear model 2) with task-based effector ROI.**

| | FC | | |
|---|---|---|---|
| *Predictors* | *Estimates* | *CI* | *p* |
| (Intercept) | 0.11 | 0.01 – 0.20 | **0.027** |
| age | -0.00 | -0.00 – 0.00 | 0.328 |
| sex [M] | 0.01 | -0.01 – 0.04 | 0.351 |
| Mean Relative Motion | -0.40 | -0.59 – -0.21 | **<0.001** |
| symptom duration | -0.00 | -0.00 – -0.00 | **0.001** |
| group [LD] | 0.03 | -0.08 – 0.14 | 0.607 |
| group [FHD] | -0.01 | -0.12 – 0.11 | 0.891 |
| ROIpair [SCAN-THa] | -0.00 | -0.11 – 0.11 | 0.999 |
| ROIpair [SCAN-Putm] | 0.11 | -0.00 – 0.21 | 0.055 |
| ROIpair [SCAN-smCB] | -0.12 | -0.23 – -0.02 | **0.024** |
| ROIpair [Foot-THp] | -0.02 | -0.13 – 0.08 | 0.673 |
| ROIpair [Foot-THa] | -0.08 | -0.19 – 0.03 | 0.141 |
| ROIpair [Foot-Putm] | -0.01 | -0.12 – 0.09 | 0.813 |
| ROIpair [Foot-smCB] | -0.09 | -0.20 – 0.01 | 0.083 |
| ROIpair [TaskHand-THp] | -0.06 | -0.16 – 0.05 | 0.309 |
| ROIpair [TaskHand-THa] | -0.17 | -0.28 – -0.07 | **0.002** |
| ROIpair [TaskHand-Putm] | -0.13 | -0.24 – -0.02 | **0.019** |
| ROIpair [TaskHand-smCB] | -0.11 | -0.22 – -0.00 | **0.049** |
| ROIpair [TaskVocal-THp] | -0.05 | -0.16 – 0.06 | 0.377 |
| ROIpair [TaskVocal-THa] | -0.07 | -0.18 – 0.03 | 0.180 |
| ROIpair [TaskVocal-Putm] | 0.05 | -0.05 – 0.16 | 0.320 |
| ROIpair [TaskVocal-smCB] | -0.17 | -0.28 – -0.06 | **0.002** |
| group [LD] × ROIpair [SCAN-THa] | 0.01 | -0.14 – 0.16 | 0.917 |
| group [FHD] × ROIpair [SCAN-THa] | -0.06 | -0.22 – 0.10 | 0.434 |
| group [LD] × ROIpair [SCAN-Putm] | 0.03 | -0.12 – 0.18 | 0.729 |
| group [FHD] × ROIpair [SCAN-Putm] | 0.04 | -0.12 – 0.20 | 0.610 |
| group [LD] × ROIpair [SCAN-smCB] | 0.12 | -0.03 – 0.27 | <u>0.111</u> |
| group [FHD] × ROIpair [SCAN-smCB] | 0.14 | -0.02 – 0.30 | <u>0.088</u> |
| group [LD] × ROIpair [Foot-THp] | -0.01 | -0.16 – 0.14 | 0.886 |
| group [FHD] × ROIpair [Foot-THp] | -0.01 | -0.17 – 0.15 | 0.918 |
| group [LD] × ROIpair [Foot-THa] | -0.02 | -0.17 – 0.13 | 0.819 |
| group [FHD] × ROIpair [Foot-THa] | -0.08 | -0.24 – 0.08 | 0.340 |
| group [LD] × ROIpair [Foot-Putm] | -0.02 | -0.17 – 0.13 | 0.754 |
| group [FHD] × ROIpair [Foot-Putm] | 0.01 | -0.15 – 0.17 | 0.924 |

| | | | |
|---|---|---|---|
| group [LD] × ROIpair [Foot-smCB] | -0.01 | -0.16 – 0.14 | 0.913 |
| group [FHD] × ROIpair [Foot-smCB] | 0.08 | -0.08 – 0.24 | 0.332 |
| group [LD] × ROIpair [TaskHand-THp] | 0.01 | -0.14 – 0.16 | 0.918 |
| group [FHD] × ROIpair [TaskHand-THp] | 0.04 | -0.12 – 0.20 | 0.618 |
| group [LD] × ROIpair [TaskHand-THa] | -0.00 | -0.15 – 0.15 | 0.965 |
| group [FHD] × ROIpair [TaskHand-THa] | -0.04 | -0.20 – 0.12 | 0.596 |
| group [LD] × ROIpair [TaskHand-Putm] | 0.01 | -0.14 – 0.16 | 0.930 |
| group [FHD] × ROIpair [TaskHand-Putm] | 0.08 | -0.08 – 0.24 | 0.326 |
| group [LD] × ROIpair [TaskHand-smCB] | 0.05 | -0.10 – 0.20 | 0.545 |
| group [FHD] × ROIpair [TaskHand-smCB] | 0.09 | -0.07 – 0.25 | 0.258 |
| group [LD] × ROIpair [TaskVocal-THp] | 0.04 | -0.11 – 0.19 | 0.615 |
| group [FHD] × ROIpair [TaskVocal-THp] | 0.04 | -0.12 – 0.20 | 0.644 |
| group [LD] × ROIpair [TaskVocal-THa] | 0.05 | -0.10 – 0.20 | 0.518 |
| group [FHD] × ROIpair [TaskVocal-THa] | -0.01 | -0.17 – 0.14 | 0.854 |
| group [LD] × ROIpair [TaskVocal-Putm] | 0.02 | -0.13 – 0.17 | 0.838 |
| group [FHD] × ROIpair [TaskVocal-Putm] | 0.07 | -0.09 – 0.23 | 0.385 |
| group [LD] × ROIpair [TaskVocal-smCB] | 0.12 | -0.03 – 0.27 | 0.117 |
| group [FHD] × ROIpair [TaskVocal-smCB] | 0.12 | -0.03 – 0.28 | 0.122 |
| Observations | 976 | | |
| $R^2$ / $R^2$ adjusted | 0.195 / 0.150 | | |

**Supplementary Table 4: Cortico-subcortical-cerebellar FC (linear model 2) with resting-state effector ROI.**

| | FC | | |
|---|---|---|---|
| *Predictors* | *Estimates* | *CI* | *p* |
| (Intercept) | 0.11 | 0.01 – 0.21 | **0.028** |
| age | -0.00 | -0.00 – 0.00 | 0.362 |
| sex [M] | 0.01 | -0.02 – 0.03 | 0.626 |
| Mean Relative Motion | -0.39 | -0.58 – -0.19 | **<0.001** |
| symptom duration | -0.00 | -0.00 – -0.00 | **<0.001** |
| group [LD] | 0.03 | -0.08 – 0.14 | 0.600 |
| group [FHD] | -0.00 | -0.12 – 0.11 | 0.947 |
| ROIpair [SCAN-THa] | -0.00 | -0.11 – 0.11 | 0.999 |
| ROIpair [SCAN-Putm] | 0.11 | -0.00 – 0.21 | 0.061 |
| ROIpair [SCAN-smCB] | -0.12 | -0.23 – -0.01 | **0.027** |
| ROIpair [Foot-THp] | -0.02 | -0.13 – 0.09 | 0.680 |
| ROIpair [Foot-THa] | -0.08 | -0.19 – 0.03 | 0.150 |
| ROIpair [Foot-Putm] | -0.01 | -0.12 – 0.10 | 0.817 |
| ROIpair [Foot-smCB] | -0.09 | -0.20 – 0.02 | 0.090 |
| ROIpair [RestHand-THp] | -0.07 | -0.18 – 0.04 | 0.199 |
| ROIpair [RestHand-THa] | -0.19 | -0.30 – -0.08 | **0.001** |
| ROIpair [RestHand-Putm] | -0.12 | -0.23 – -0.01 | **0.028** |
| ROIpair [RestHand-smCB] | -0.10 | -0.21 – 0.01 | 0.066 |
| ROIpair [RestMouth-THp] | -0.01 | -0.12 – 0.10 | 0.849 |
| ROIpair [RestMouth-THa] | -0.09 | -0.20 – 0.02 | 0.108 |
| ROIpair [RestMouth-Putm] | -0.02 | -0.13 – 0.09 | 0.753 |
| ROIpair [RestMouth-smCB] | -0.17 | -0.28 – -0.06 | **0.002** |
| group [LD] × ROIpair [SCAN-THa] | 0.01 | -0.15 – 0.16 | 0.919 |
| group [FHD] × ROIpair [SCAN-THa] | -0.06 | -0.22 – 0.10 | 0.444 |
| group [LD] × ROIpair [SCAN-Putm] | 0.03 | -0.13 – 0.18 | 0.735 |
| group [FHD] × ROIpair [SCAN-Putm] | 0.04 | -0.12 – 0.20 | 0.618 |
| group [LD] × ROIpair [SCAN-smCB] | 0.12 | -0.03 – 0.28 | 0.120 |
| group [FHD] × ROIpair [SCAN-smCB] | 0.14 | -0.02 – 0.30 | 0.095 |
| group [LD] × ROIpair [Foot-THp] | -0.01 | -0.16 – 0.14 | 0.888 |
| group [FHD] × ROIpair [Foot-THp] | -0.01 | -0.17 – 0.15 | 0.920 |
| group [LD] × ROIpair [Foot-THa] | -0.02 | -0.17 – 0.14 | 0.823 |
| group [FHD] × ROIpair [Foot-THa] | -0.08 | -0.24 – 0.08 | 0.351 |
| group [LD] × ROIpair [Foot-Putm] | -0.02 | -0.18 – 0.13 | 0.759 |
| group [FHD] × ROIpair [Foot-Putm] | 0.01 | -0.15 – 0.17 | 0.926 |

| | | | |
|---|---|---|---|
| group [LD] × ROIpair [Foot-smCB] | -0.01 | -0.16 – 0.15 | 0.915 |
| group [FHD] × ROIpair [Foot-smCB] | 0.08 | -0.08 – 0.24 | 0.343 |
| group [LD] × ROIpair [RestHand-THp] | 0.01 | -0.14 – 0.16 | 0.909 |
| group [FHD] × ROIpair [RestHand-THp] | 0.03 | -0.13 – 0.19 | 0.698 |
| group [LD] × ROIpair [RestHand-THa] | -0.03 | -0.18 – 0.12 | 0.706 |
| group [FHD] × ROIpair [RestHand-THa] | -0.05 | -0.21 – 0.11 | 0.559 |
| group [LD] × ROIpair [RestHand-Putm] | 0.01 | -0.14 – 0.16 | 0.907 |
| group [FHD] × ROIpair [RestHand-Putm] | 0.06 | -0.10 – 0.23 | 0.441 |
| group [LD] × ROIpair [RestHand-smCB] | 0.02 | -0.14 – 0.17 | 0.838 |
| group [FHD] × ROIpair [RestHand-smCB] | 0.08 | -0.08 – 0.25 | 0.303 |
| group [LD] × ROIpair [RestMouth-THp] | 0.02 | -0.13 – 0.18 | 0.768 |
| group [FHD] × ROIpair [RestMouth-THp] | 0.05 | -0.11 – 0.21 | 0.530 |
| group [LD] × ROIpair [RestMouth-THa] | 0.01 | -0.14 – 0.17 | 0.871 |
| group [FHD] × ROIpair [RestMouth-THa] | -0.02 | -0.19 – 0.14 | 0.765 |
| group [LD] × ROIpair [RestMouth-Putm] | -0.01 | -0.16 – 0.15 | 0.945 |
| group [FHD] × ROIpair [RestMouth-Putm] | 0.06 | -0.10 – 0.22 | 0.480 |
| group [LD] × ROIpair [RestMouth-smCB] | 0.08 | -0.07 – 0.24 | 0.284 |
| group [FHD] × ROIpair [RestMouth-smCB] | 0.12 | -0.04 – 0.28 | 0.155 |
| Observations | 976 | | |
| $R^2$ / $R^2$ adjusted | 0.188 / 0.144 | | |

### Supplementary Table 5: Subcortical-cerebellar FC (linear model 3).

| | FC | | |
|---|---|---|---|
| *Predictors* | *Estimates* | *CI* | *p* |
| (Intercept) | 0.83 | 0.73 – 0.93 | **<0.001** |
| age | -0.00 | -0.00 – 0.00 | 0.125 |
| sex [M] | 0.02 | -0.02 – 0.05 | 0.324 |
| Mean Relative Motion | 0.53 | 0.29 – 0.78 | **<0.001** |
| symptom duration | 0.00 | 0.00 – 0.00 | **0.047** |
| group [LD] | -0.01 | -0.12 – 0.09 | 0.823 |
| group [FHD] | -0.03 | -0.14 – 0.08 | 0.626 |
| ROIpair [GPe-STN] | -0.28 | -0.39 – -0.18 | **<0.001** |
| ROIpair [GPi-STN] | -0.26 | -0.36 – -0.15 | **<0.001** |
| ROIpair [GPi-THa] | -0.52 | -0.62 – -0.41 | **<0.001** |
| ROIpair [GPi-THp] | -0.57 | -0.67 – -0.46 | **<0.001** |
| ROIpair [Putm-GPi] | -0.44 | -0.55 – -0.34 | **<0.001** |
| ROIpair [Putm-Caud] | -0.06 | -0.17 – 0.04 | 0.235 |
| ROIpair [smCB-THa] | -0.72 | -0.83 – -0.62 | **<0.001** |
| ROIpair [smCB-THp] | -0.80 | -0.90 – -0.69 | **<0.001** |
| group [LD] × ROIpair [GPe-STN] | 0.02 | -0.12 – 0.16 | 0.782 |
| group [FHD] × ROIpair [GPe-STN] | -0.03 | -0.18 – 0.12 | 0.717 |
| group [LD] × ROIpair [GPi-STN] | 0.00 | -0.14 – 0.14 | 0.999 |
| group [FHD] × ROIpair [GPi-STN] | -0.03 | -0.18 – 0.13 | 0.737 |
| group [LD] × ROIpair [GPi-THa] | -0.12 | -0.26 – 0.03 | 0.116 |
| group [FHD] × ROIpair [GPi-THa] | -0.03 | -0.18 – 0.12 | 0.666 |
| group [LD] × ROIpair [GPi-THp] | -0.09 | -0.24 – 0.05 | 0.211 |
| group [FHD] × ROIpair [GPi-THp] | -0.06 | -0.21 – 0.09 | 0.437 |
| group [LD] × ROIpair [Putm-GPi] | -0.05 | -0.19 – 0.10 | 0.518 |
| group [FHD] × ROIpair [Putm-GPi] | -0.01 | -0.16 – 0.14 | 0.891 |
| group [LD] × ROIpair [Putm-Caud] | 0.01 | -0.14 – 0.15 | 0.916 |
| group [FHD] × ROIpair [Putm-Caud] | -0.05 | -0.20 – 0.10 | 0.515 |
| group [LD] × ROIpair [smCB-THa] | -0.04 | -0.18 – 0.10 | 0.577 |
| group [FHD] × ROIpair [smCB-THa] | -0.03 | -0.18 – 0.12 | 0.684 |
| group [LD] × ROIpair [smCB-THp] | 0.01 | -0.13 – 0.16 | 0.876 |
| group [FHD] × ROIpair [smCB-THp] | -0.04 | -0.19 – 0.11 | 0.613 |
| Observations | 549 | | |
| $R^2$ / $R^2$ adjusted | 0.733 / 0.717 | | |

### Supplementary Table 6: SCAN-to-sensorimotor-cerebellum linear model with combined dystonia group.

| | FC | | |
|---|---|---|---|
| *Predictors* | *Estimates* | *CI* | *p* |
| (Intercept) | 0.10 | -0.15 – 0.34 | 0.426 |
| group2 [Dystonia] | 0.18 | 0.05 – 0.32 | **0.010** |
| age | -0.00 | -0.01 – 0.00 | 0.443 |
| sex [M] | 0.02 | -0.08 – 0.12 | 0.647 |
| symptom duration | -0.00 | -0.01 – 0.00 | 0.182 |
| Mean Relative Motion | -0.97 | -1.76 – -0.18 | **0.017** |
| Observations | 61 | | |
| $R^2$ / $R^2$ adjusted | 0.204 / 0.132 | | |
